# Effectiveness of the BNT162b2 vaccine among children 5-11 and 12-17 years in New York after the Emergence of the Omicron Variant

**DOI:** 10.1101/2022.02.25.22271454

**Authors:** Vajeera Dorabawila, Dina Hoefer, Ursula E. Bauer, Mary T. Bassett, Emily Lutterloh, Eli S. Rosenberg

## Abstract

**Importance:** There is limited evidence on the effectiveness of the BNT162b2 vaccine for children, particularly those 5-11 years and after the Omicron variant’s emergence.

**Objective:** To estimate BNT162b2 vaccine effectiveness against COVID cases and hospitalizations among children 5-11 years and 12-17 years during December, 2021 and January, 2022.

**Design:** Analyses of cohorts constructed from linked statewide immunization, laboratory testing, and hospitalization databases.

**Setting/Participants:** New York State children 5-17 years.

**Main outcomes/measures:** New laboratory-confirmed COVID-19 cases and hospitalizations. Comparisons were made using the incidence rate ratio (IRR), comparing outcomes by vaccination status, and estimated vaccine effectiveness (VE: 1-[1/IRR]).

**Results:** From December 13, 2021 to January 30, 2022, among 852,384 fully-vaccinated children 12-17 years and 365,502 children 5-11 years, VE against cases declined from 66% (95% CI: 64%, 67%) to 51% (95% CI: 48%, 54%) for those 12-17 years and from 68% (95% CI: 63%, 72%) to 12% (95% CI: 6%, 16%) for those 5-11 years. During the January 24-30 week, VE for children 11 years was 11% (95%CI -3%, 23%) and for those age 12 was 67% (95% CI: 62%, 71%). VE against hospitalization declined changed from 85% (95% CI: 63%, 95%) to 73% (95% CI: 53%, 87%) for children 12-17 years, and from 100% (95% CI: -189%, 100%) to 48% (95% CI: -12%, 75%) for those 5-11 years. Among children newly fully-vaccinated December 13, 2021 to January 2, 2022, VE against cases within two weeks of full vaccination for children 12-17 years was 76% (95% CI: 71%, 81%) and by 28-34 days it was 56% (95% CI: 43%, 63%). For children 5-11, VE against cases declined from 65% (95% CI: 62%, 68%) to 12% (95% CI: 8%, 16%) by 28-34 days.

**Conclusions and Relevance:** In the Omicron era, the effectiveness against cases of BNT162b2 declined rapidly for children, particularly those 5-11 years. However, vaccination of children 5-11 years was protective against severe disease and is recommended. These results highlight the potential need to study alternative vaccine dosing for children and the continued importance layered protections, including mask wearing, to prevent infection and transmission.

## Introduction

In New York State (NYS), nearly 850,000 children ≤17 years have been diagnosed with COVID-19. Randomized trails and observational studies conducted during the Delta and earlier variants’ predominance, indicate the BNT162b2 vaccine, developed to protect against original strains, is safe and effective in preventing COVID-19 outcomes in those 5-17 years and older.^1-10^ Compared to children 12-17 years, who receive two 30μg doses, less is known known about real-world vaccine effectiveness against infection and hospitalization effectiveness for children 5-11 years, who receive two 10μg doses, particularly after the Omicron variant’s emergence.^11,12^ We examined the effectiveness of vaccination during the Omicron variant surge that began in early December 2021 on infection and hospitalization among children 5-11 years compared to 12-17 years using NYS statewide surveillance systems.

## Methods

Three NYS databases were linked to examine COVID-19 outcomes among children 5-17 years.^6,7,8^ The Citywide Immunization Registry (CIR) and the NYS Immunization Information System (NYSIIS) collect COVID-19 provider vaccination data for residents of New York City and the rest of NYS, respectively. The Electronic Clinical Laboratory Reporting System (ECLRS) collects all reportable COVID-19 test results. The Health Electronic Response Data System (HERDS) includes a statewide, daily electronic survey of inpatient facilities, including all new admissions with laboratory-confirmed COVID-19 and the primary reason for admission.

The analysis compared outcomes among fully vaccinated children (defined as series completion + 14 days) versus unvaccinated children in the age groups 5-11 years and 12 -17 years. Two outcomes were assessed: 1) COVID-19 cases, defined as positive NAAT or antigen results reported to ECLRS, 2) New COVID-19 admissions as reported in HERDS.

Cases and admissions were enumerated weekly for the open cohorts of vaccinated and unvaccinated (*Census population minus partially and fully vaccinated populations*) children 5-11 and 12-17 years, from November 29, 2021 to January 30, 2022.^7,8^ Weekly rates of new cases and hospitalizations were calculated by respectively dividing each group count by the fully vaccinated and unvaccinated person-days in that week. Within each age group, incidence rate ratios (IRR) were calculated by dividing the unvaccinated rate by the vaccinated rate, and vaccine effectiveness (VE) calculated as 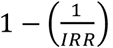, with 95% confidence intervals. To probe the potential role of vaccine dose, the case analysis was repeated by single year age at vaccination.

Additionally, limiting analysis to children newly fully-vaccinated in the 3 weeks from December 13, 2021 to January 2, 2022 permitted examination of time since vaccination. Case rates for the 3 weekly cohorts were estimated by time-since-vaccination across the January 3-30, 2022 period, and compared to those contemporaneously unvaccinated using IRR and VE, separately for those 5-11 and 12-17 years (eTable 1, Supplemental Methods).

**Table 1:** New COVID-19 Cases and Hospitalizations by Vaccine Status, Children Ages 5-17 in New York State, November 29, 2021 – January 30, 2022.

|  | Events |  | Rates per 100k |  | Incidence Rate Ratios, Vaccine Effectiveness |  |  |  | Full-vaccine Coverage |
| --- | --- | --- | --- | --- | --- | --- | --- | --- | --- |
| Week | Vacci-nated | Unvacci-nated | Vacci-nated | Unvacci-nated | IRR | (95% CI) | VE | (95% CI) | % |
| **Cases** |  |  |  |  |  |  |  |  |  |
| **5 -11 years<sup>a</sup>** |  |  |  |  |  |  |  |  |  |
| Dec. 13-19 | 204 | 10,062 | 39 | 122 | 3.1 | (2.7, 3.6) | 68% | (63, 72%) | 4.7% |
| Dec. 20-26 | 1,051 | 16,915 | 91 | 212 | 2.3 | (2.2, 2.5) | 57% | (55, 60%) | 10.6% |
| Dec. 27-Jan. 2 | 2,910 | 28,476 | 184 | 369 | 2.0 | (1.9, 2.1) | 50% | (48, 52%) | 14.4% |
| Jan. 3-9 | 4,535 | 36,256 | 251 | 484 | 1.9 | (1.9, 2.0) | 48% | (47, 50%) | 16.5% |
| Jan. 10-16 | 3,459 | 18,647 | 170 | 256 | 1.5 | (1.5, 1.6) | 34% | (31, 36%) | 18.5% |
| Jan. 17-23 | 2,403 | 9,289 | 105 | 131 | 1.3 | (1.2, 1.3) | 20% | (16, 23%) | 20.9% |
| Jan. 24-30 | 1,584 | 4,889 | 62 | 70 | 1.1 | (1.1, 1.2) | 12% | (6, 16%) | 23.4% |
| **12 – 17 years** |  |  |  |  |  |  |  |  |  |
| Nov. 29-Dec. 5 | 848 | 3,336 | 15 | 101 | 6.7 | (6.2, 7.2) | 85% | (84, 86%) | 58.5% |
| Dec. 6-12 | 1,302 | 4,245 | 23 | 131 | 5.7 | (5.3, 6.0) | 82% | (81, 83%) | 58.9% |
| Dec. 13-19 | 4,140 | 6,738 | 73 | 212 | 2.9 | (2.8, 3.0) | 66% | (64, 67%) | 59.4% |
| Dec. 20-26 | 10,177 | 12,792 | 178 | 411 | 2.3 | (2.3, 2.4) | 57% | (56, 58%) | 59.9% |
| Dec. 27-Jan. 2 | 15,934 | 18,905 | 276 | 617 | 2.2 | (2.2, 2.3) | 55% | (54, 56%) | 60.5% |
| Jan. 3-9 | 19,219 | 21,333 | 331 | 706 | 2.1 | (2.1, 2.2) | 53% | (52, 54%) | 60.9% |
| Jan. 10-16 | 10,185 | 10,264 | 174 | 345 | 2.0 | (1.9, 2.0) | 50% | (48, 51%) | 61.4% |
| Jan. 17-23 | 5,010 | 4,971 | 85 | 169 | 2.0 | (1.9, 2.1) | 50% | (48, 52%) | 62.0% |
| Jan. 24-30 | 2,581 | 2,575 | 43 | 88 | 2.0 | (1.9, 2.2) | 51% | (48, 54%) | 62.5% |
| **Hospitalizations** |  |  |  |  |  |  |  |  |  |
| **5 -11 years<sup>a</sup>** |  |  |  |  |  |  |  |  |  |
| Dec. 13-19 | 0 | 18 | 0.00 | 0.22 | +inf. | (0.3, +inf.) | 100% | (-189, 100%) | 4.7% |
| Dec. 20-26 | 2 | 50 | 0.17 | 0.63 | 3.6 | (1.0, 30.9) | 73% | (-7, 97%) | 10.6% |
| Dec. 27-Jan. 2 | 3 | 80 | 0.19 | 1.04 | 5.5 | (1.8, 27.1) | 82% | (45, 96%) | 14.5% |
| Jan. 3-9 | 5 | 78 | 0.28 | 1.04 | 3.8 | (1.6, 12.0) | 74% | (36, 96%) | 16.6% |
| Jan. 10-16 | 6 | 68 | 0.29 | 0.94 | 3.2 | (1.4, 8.9) | 68% | (28, 91%) | 18.6% |
| Jan. 17-23 | 8 | 46 | 0.35 | 0.65 | 1.9 | (0.9, 4.6) | 46% | (-15, 77%) | 21.0% |
| Jan. 24-30 | 8 | 42 | 0.31 | 0.60 | 1.9 | (0.9, 4.8) | 48% | (-12, 75%) | 23.4% |
| **12 – 17 years** |  |  |  |  |  |  |  |  |  |
| Nov. 29-Dec. 5 | 2 | 20 | 0.04 | 0.61 | 16.9 | (4.1, 148.8) | 94% | (76, 99%) | 58.4% |
| Dec. 6-12 | 1 | 11 | 0.02 | 0.34 | 19.0 | (2.8, 818.3) | 95% | (64, 100%) | 58.8% |
| Dec. 13-19 | 6 | 23 | 0.11 | 0.72 | 6.8 | (2.7, 20.4) | 85% | (63, 95%) | 59.3% |
| Dec. 20-26 | 18 | 45 | 0.31 | 1.44 | 4.6 | (2.6, 8.4) | 78% | (63, 88%) | 59.9% |
| Dec. 27-Jan. 2 | 38 | 77 | 0.66 | 2.50 | 3.8 | (2.5, 5.8) | 74% | (61, 84%) | 60.4% |
| Jan. 3-9 | 47 | 94 | 0.81 | 3.10 | 3.8 | (2.7, 5.6) | 74% | (63, 82%) | 60.9% |
| Jan. 10-16 | 41 | 85 | 0.70 | 2.84 | 4.1 | (2.8, 6.0) | 75% | (64, 86%) | 61.3% |
| Jan. 17-23 | 34 | 67 | 0.58 | 2.26 | 3.9 | (2.6, 6.1) | 75% | (61, 83%) | 61.9% |
| Jan. 24-30 | 22 | 40 | 0.37 | 1.36 | 3.7 | (2.1, 6.5) | 73% | (53, 87%) | 62.4% |
| <sup>a</sup> <1% of this age group fully vaccinated in previous weeks |  |  |  |  |  |  |  |  |  |

The New York State Department of Health Institutional Review Board (IRB) determined this is a surveillance activity necessary for public health work, and therefore waived the need for IRB review.

## Results

By the week of January 24-30, 2022, 365,502 children 5-11 (23.4%) and 852,384 children 12-17 years (62.4%) were fully-vaccinated in NYS. Within age groups, there was little difference in mean age by vaccination status (5-11: 8.3 vs. 7.8 years, 12-17: 14.6 vs 14.6 years). The median time since vaccination was respectively 51 and 211 days for children 5-11 and 12-17 years (eTable 2).

For children 12-17 years, the unvaccinated vs. vaccinated IRR for cases declined from 6.7 (95% CI: 6.2, 7.2; VE: 85% [95% CI: 84%, 86%]) the week of November 29 to 2.9 (95% CI: 2.8, 3.0; VE: 66% [95% CI: 64%, 67%]) the week of December 13, when the Omicron variant was 19% of sequences,^13^ and declined further to 2.0 (95% CI: 1.9, 2.2; VE: 51% [95% CI: 48%, 54%]) by January 24, when Omicron exceeded 99% of sequences (Table 1). For those 5-11 years, during the December 13 week IRR was 3.1 (95% CI: 2.7, 3.6; VE: 68% [95% CI: 63%, 72%]), but declined to 1.1 (95% CI: 1.1, 1.2; VE: 12% [95% CI: 6%, 16%]) by January 24. Compared to cases, protection was higher for hospitalization by January 24, with IRR 1.9 (95% CI: 0.9, 4.8; VE: 48% [95% CI: -12%, 75%] for 5-11 years and IRR 3.7 (95% CI: 2.1,6.5; VE: 73% [95% CI: 53%, 87%]) for 12-17 years although outcomes were sparser, particularly when restricted to primary admission reason for COVID-19 (eTable 3).

By single-year age, all 5–11-year VE estimates against infection declined to below 12-17-year estimates by January 24 (Figure 1), with no clear age gradient. That week, VE was 11% (95% CI: -3%, 23%) for age 11 versus 67% (95% CI: 62%, 71%) for age 12.

**Figure 1:**
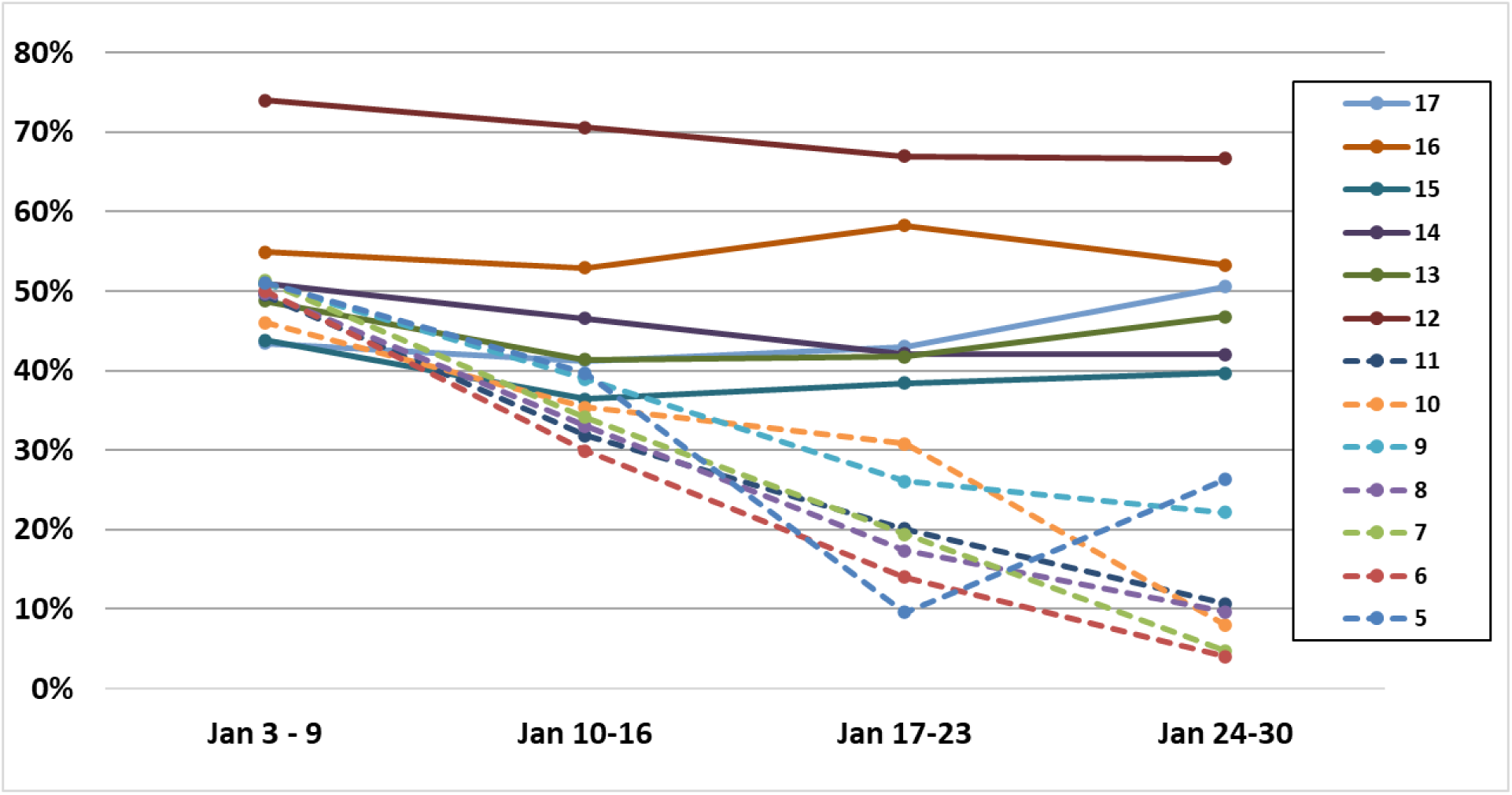
Vaccine Effectiveness against Infection, by Week and Year of Age.

Examining time-since-vaccination, ≤13 days of full-vaccination, 12-17 years IRR was 4.3 (95% CI: 3.4, 5.3; VE: 76% [95% CI: 71%, 81%]), but by 28-34 days it was 2.3 (95% CI: 1.9, 2.7; VE: 56% [95% CI: 48%, 63%], Figure 2). For children 5-11 years, IRR at ≤13 days was 2.9 (95% CI 2.7, 3.1; VE: 65% [95% CI: 62%, 68%]) and at 28-34 days it was 1.1 (95% CI: 1.1, 1.2; VE: 12% [95% CI: 8%, 16%]).

**Figure 2:**
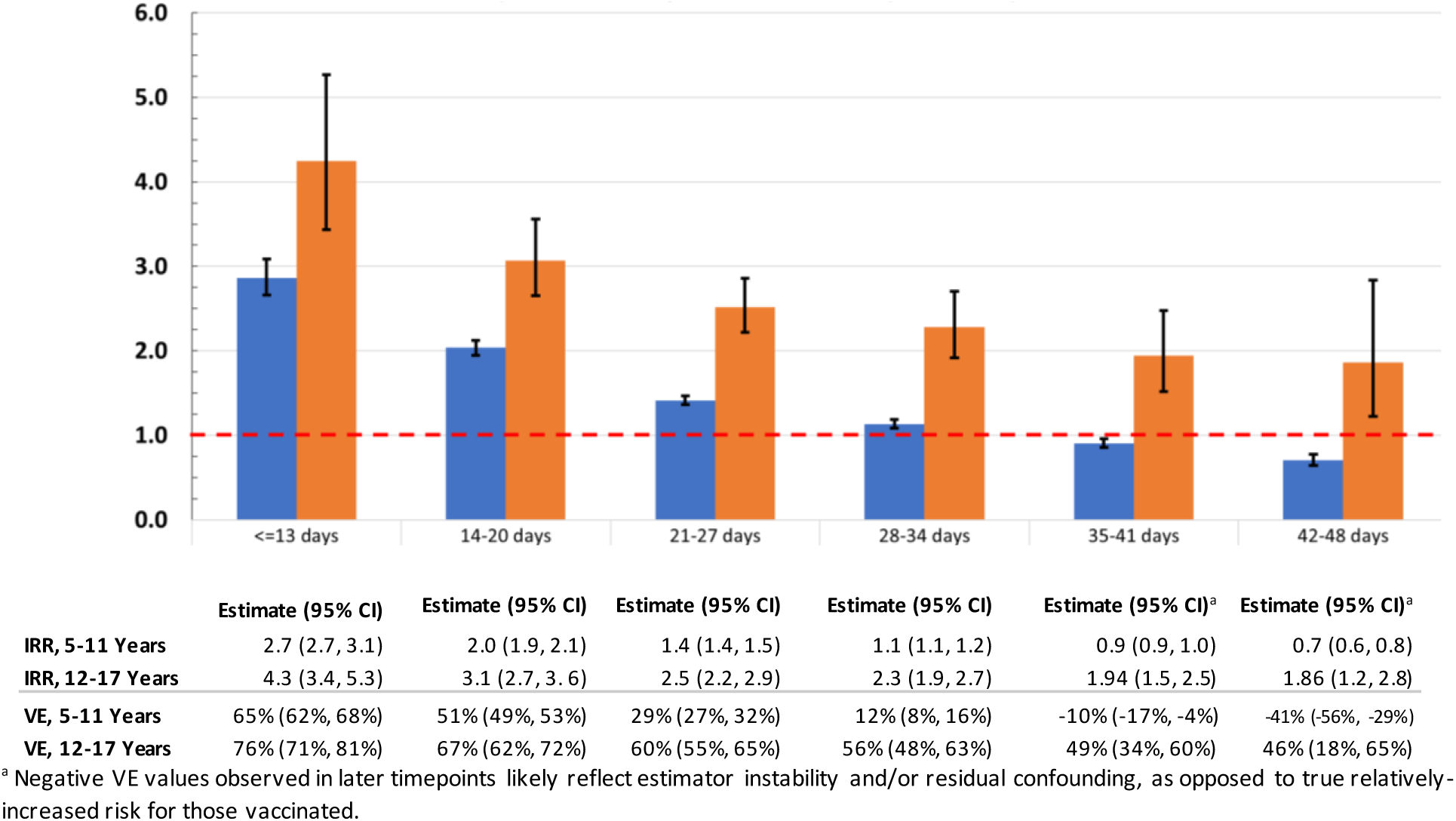
Incidence rate ratios, comparing cases during January 3 - January 30, 2022 for unvaccinated versus children newly fully-vaccinated December 13, 2021-January 2, 2022, by Time Since Full Vaccination.

## Discussion

During Omicron variant predominance, VE against infection declined rapidly for NYS children 5-11 years, with low protection by one month following full-vaccination. Among children 12-17, protection declined substantially, albeit more slowly than observed among younger children. These results complement early findings of reduced primary series VE for ≥12 years during the Omicron era and the dual effects of the variant and waning protection against infection with sustained protection against severe disease.^1,8,10^

The finding of markedly-lower VE against infection for children 11 years compared to those 12 and 13 years, despite overlapping physiology, suggests lower vaccine dose may explain lower 5-11 years VE. Children 12 years had the highest VE of all ages, potentially due to being small size relative to dose and more recent vaccination (by 6 weeks on average) than those 13-17 years. This gap suggests a threshold effect between the two BNT162b2 vaccine doses and need for study of numbers of doses, amount per dose, dose timing, and/or antigens targeted for children 5-11 years.

The findings in this study are subject to limitations. First, home testing, whichis not reported and increased during the analysis period, would impact conclusions if testing practices differed by vaccine status. This potential bias would not impact hospitalizations. During this highest-incidence period, including for severe disease, for children, there were still relatively few children admissions. Additional data are needed to fully-understand VE trends against severe disease.^2,9^ By the end of January, 12.5% of vaccinated children 12-17 years had received a booster, likely adding protection to that group, although the time-since-vaccination analysis had no boosted children. This analysis compared early vaccinators in the younger age group withlater vaccinators in the older age group, who may differ in test-seeking or exposures. Similar conclusions regarding relative VE declines between children 5-11 and 12-17 years using open cohorts suggests this trend is robust.

Our data support vaccine protection against severe disease among children 5-11 years, but suggest rapid loss of protection against infection, in the Omicron variant era. Should such findings be replicated in other settings, review of the dosing schedule for children 5-11 years appears prudent. At this time, efforts to increase primary vaccination coverage in this age group, which remains <25% nationally, should continue. Given rapid loss of protection against infections, these results highlight the continued importance of layered protections, including mask wearing, for children to prevent infection and transmission.

## Data Availability

All data produced in the present study are available upon reasonable request to the authors

