## Supplement for "Effectiveness of the BNT162b2 vaccine among children 5-11 and 12-17 years in New York after the Emergence of the Omicron Variant"

**eMethods**

**eTable 1.** New Cases among Children Newly Vaccinated from December 13, 2021 to January 2, 2022

**eTable 2.** Days Since Series Completion by Age as of January 3, 2022 and January 31, 2022

**eTable 3.** New Hospitalizations with Primary Reason for Admissions being for COVID-19, Incidence Rate Ratio, and Vaccine Effectiveness, Children Ages 5-17

**eTable 4.** Time since vaccination analysis: Rates per 100,000, Incidence Rate Ratio, and Vaccine Effectiveness

### eMethods

#### Dataset construction

Vaccination registry (NYSIIS/CIR) COVID-19 data were combined and deduplicated based on first name, last name, date of birth (DOB), and ZIP code then matched to laboratory data (ECLRS) using a deterministic algorithm based on first name, last name, and DOB, and to hospital admissions (HERDS) based on initials, sex, DOB, and ZIP code. Partially and fully vaccinated children in each age group were subtracted from the census population in the applicable age groups to arrive at the unvaccinated children count. Partially vaccinated children were excluded from the analyses.

#### Time since vaccination analysis

This analysis included three weekly cohorts of children who completed the primary series from December 13, 2021 to January 2, 2022. There were 1,539,762 person days of children aged 5-11 years and 151,005 person days of children aged 12-17 years (eTable 1). The number of children aged 5-11 years included in this analysis was higher compared those 12-17 years, largely as a function of when the vaccine was authorized for these age groups. US Food and Drug Administration emergency use authorization for children 16-17 years was given on December 11, 2020, 12-15 years on May 10, 2021 and aged 5-11 years on October 29, 2021.

Outcomes among the three weekly cohorts were arranged into time-since-vaccination categories during the four January 3-30, 2022 follow-up weeks by combining cohorts as illustrated via colored diagonal cells in eTable 1. For example, cases within ≤13 days of full-vaccination (grey shading) are children that were vaccinated between December 27-January 2 with an outcome window between January 3-9, 2022. Similarly, cases 28-34 days after full-vaccination (light-blue shading) represented the sum of the following 3 groups: (1) persons vaccinated between December 12-19 and with cases between January 10-16 (1,234 cases in the 5-11 age group, for example); (2) persons vaccinated between December 20-26 and with cases between January 17-23 (635 cases in the 5-11 age group); and (3) persons vaccinated between December 27-January 2 and with cases between January 24-30 (204 cases in the 5-11 age group).

Incident case rates among the vaccinated, by time-since-vaccination interval, were constructed. The numerator, cases, was the sum in each time-since-vaccination group (2,073 in the example of cases between 28-34 days in the 5-11 age group). In this example, there were 3 vaccination groups (light blue) that were combined, per the colored diagonals in eTable 1. The denominator is the number of children observed fully-vaccinated during the time-since-vaccination interval and contributing to the numerator. In the example of 28-34 days, all three weekly cohorts were vaccinated for that interval, thus 1,539,762 person days of children 5-11 years contributed to the denominator. Thus incidence rates for those vaccinated ($R_V$) were $R_V = \frac{\sum_{i=1}^{k} C_{V,i}}{\sum_{i=1}^{k} N_{V,i}}$, with $var(R_V) = \frac{\sum_{i=1}^{k} C_{V,i}}{\left(\sum_{i=1}^{k} N_{V,i}\right)^2}$, where $i$ is the cohort, $k$ is the number of cohorts combined, $C_{V,i}$ is the number of cases among vaccinated cohort members, and $N_{V,i}$ is the number of vaccinated person days observed in the cohort for the interval.[1]

Rates among an unvaccinated comparison group were constructed from unvaccinated rates in the same weeks contributing to a given time-since-vaccination interval, directly standardized to the person-time of those vaccinated, as follows. The directly-standardized rate for the unvaccinated ($DSR_U$), was estimated as $DSR_U = \frac{\sum_{i=1}^{k} N_{V,i} R_{U,i}}{\sum_{i=1}^{k} N_{V,i}}$, with $var(DSR_U) = \frac{\sum_{i=1}^{k} \frac{\left(N_{V,i}\right)^2 R_{U,i}}{N_{U,i}}}{\left(\sum_{i=1}^{k} N_{V,i}\right)^2}$, where $R_{U,i}$ is the unvaccinated case rate ($\frac{C_{U,i}}{N_{U,i}}$, cases among unvaccinated / unvaccinated persons, as calculated for the open-cohort method) and $N_{U,i}$ is number of unvaccinated persons in a given week contributing to the interval (eTable 4).[2,3]

The incidence rate ratio (IRR) for each time-since-vaccination interval was estimated as $IRR = \frac{DSR_U}{R_V}$, with $var(ln[IRR]) = \frac{var(R_V)}{R_V^2} + \frac{var(DSR_U)}{DSR_U^2}$, and 95% $CI = e^{\ln(IRR) \pm 1.96\sqrt{var(\ln(IRR))}}$.[2,3]

**eTable 1: New Cases among Children Newly Vaccinated from December 13, 2021 to January 2, 2022[a]**

| Cohort | Person Days Newly Vaccinated | Follow Up Week | | | |
|---|---|---|---|---|---|
| | | Jan 3 – 9 | Jan 10-16 | Jan 17-23 | Jan 24-30 |
| **5-11 Years** | | | | | |
| Dec 13-19 | 476,265 | 1,808 | 1,234 | 781 | 472 |
| Dec 20-26 | 642,013 | 1,536 | 1,054 | 635 | 403 |
| Dec 27-Jan 2 | 421,484 | 712 | 522 | 320 | 204 |
| **Age 12-17 Years** | | | | | |
| Dec 13-19 | 46,414 | 132 | 82 | 42 | 22 |
| Dec 20-26 | 54,054 | 126 | 65 | 33 | 23 |
| Dec 27-Jan 2 | 50,537 | 84 | 55 | 41 | 15 |

[a] Days since full vaccination are indicated in background color: Grey: <=13 days, Orange: 14-20 days, Yellow: 21-27 days, Light-blue: 28-34 days, Green: 35-41 days, Dark-blue: 42-48 days

**eTable 2: Days Since Series Completion by Age as of January 3, 2022 and January 31, 2022**

| Age at Series Completion | Days from Series Completion to 1/3/22 | | | | Days from Series Completion to 1/31/22 | | | |
|---|---|---|---|---|---|---|---|---|
| | # Children | Mean | Median | SD | # Children | Mean | Median | SD |
| **5-11 Total** | 247,953 | 28 | 28 | 13 | 384,284 | 46 | 51 | 18 |
| 5 | 27,034 | 28 | 28 | 10 | 43,274 | 45 | 50 | 16 |
| 6 | 30,922 | 28 | 28 | 12 | 48,338 | 46 | 51 | 17 |
| 7 | 33,642 | 28 | 28 | 9 | 51,971 | 46 | 51 | 16 |
| 8 | 35,120 | 28 | 28 | 21 | 54,134 | 46 | 51 | 22 |
| 9 | 38,113 | 28 | 28 | 10 | 58,776 | 46 | 51 | 16 |
| 10 | 40,752 | 28 | 28 | 10 | 62,236 | 46 | 51 | 16 |
| 11 | 42,370 | 28 | 28 | 14 | 65,555 | 46 | 51 | 19 |
| **12-17 Total** | 884,233 | 168 | 187 | 60 | 914,257 | 191 | 211 | 67 |
| 12 | 146,944 | 143 | 145 | 58 | 57,061 | 162 | 165 | 66 |
| 13 | 132,238 | 160 | 177 | 51 | 136,536 | 183 | 201 | 57 |
| 14 | 39,649 | 162 | 180 | 54 | 43,504 | 185 | 205 | 60 |
| 15 | 141,304 | 163 | 185 | 51 | 45,092 | 187 | 209 | 57 |
| 16 | 159,003 | 185 | 204 | 66 | 162,944 | 208 | 231 | 71 |
| 17 | 165,095 | 191 | 211 | 64 | 169,120 | 214 | 237 | 70 |

**eTable 3: New Hospitalizations with Primary Reason for Admissions being for COVID-19, Incidence Rate Ratio, and Vaccine Effectiveness, Children Ages 5-17**

| Week | New Hospitalizations | | | | Incidence Rate Ratios, Vaccine Effectiveness | | | |
|---|---|---|---|---|---|---|---|---|
| | Vaccinated | | Unvaccinated | | | | | |
| | n | % of COVID-19 admissions | n | % of COVID-19 admissions | IRR[a] | (95% CI) | VE[a] | (95% CI) |
| **5 -11 years** | | | | | | | | |
| Dec. 13-19 | 0 | 0% | 6 | 33% | +inf. | (0.1, +inf.) | 100% | (-1256, 100%) |
| Dec. 20-26 | 2 | 100% | 29 | 58% | 2.1 | (0.5, 18.2) | 53% | (-87, 95%) |
| Dec. 27-Jan. 2 | 1 | 33% | 43 | 54% | 8.8 | (1.5, 356.0) | 89% | (33, 100%) |
| Jan. 3-9 | 3 | 60% | 31 | 40% | 2.5 | (0.8, 12.8) | 60% | (-28, 92%) |
| Jan. 10-16 | 1 | 17% | 29 | 43% | 8.1 | (1.3, 330.5) | 88% | (25, 100%) |
| Jan. 17-23 | 4 | 50% | 13 | 28% | 1.0 | (0.3, 4.4) | 4% | (-209, 77%) |
| Jan. 24-30 | 2 | 25% | 15 | 36% | 2.7 | (0.6, 24.8) | 64% | (-56, 96%) |
| **December 13 – January 30** | **13** | **41%** | **166** | **43%** | **2.9** | **(1.6, 5.5)** | **65%** | **(39, 82%** |
| **12 – 17 years** | | | | | | | | |
| Nov. 29-Dec. 5 | 0 | 0% | 8 | 40% | +inf. | (2.9, +inf.) | 100% | (65, 100%) |
| Dec. 6-12 | 0 | 0% | 6 | 55% | +inf. | (2.0, +inf) | 100% | (51, 100%) |
| Dec. 13-19 | 3 | 50% | 8 | 35% | 4.8 | (1.1, 27.9) | 79% | (13, 96%) |
| Dec. 20-26 | 5 | 28% | 19 | 42% | 7.0 | (2.5, 23.9) | 86% | (60, 96%) |
| Dec. 27-Jan. 2 | 15 | 39% | 28 | 36% | 3.5 | (1.8, 7.1) | 72% | (45, 86%) |
| Jan. 3-9 | 10 | 21% | 24 | 26% | 4.6 | (2.1, 10.8) | 78% | (53, 91%) |
| Jan. 10-16 | 9 | 22% | 23 | 27% | 5.0 | (2.2, 12.4) | 80% | (55, 92%) |
| Jan. 17-23 | 5 | 15% | 12 | 18% | 4.8 | (1.6, 17.5) | 79% | (37, 94%) |
| Jan. 24-30 | 1 | 5% | 8 | 20% | 16.4 | (2.0, 131.0) | 94% | (51, 99%) |
| **December 13 – January 30** | **48** | **23%** | **122** | **28%** | **4.9** | **(3.5, 7.0)** | **80%** | **(71, 86%)** |

[a] IRR and VE calculated on rates limited to only those COVID-19 admissions with primary reason indicated as “for COVID-19”. This represents a conservative estimate, since admissions with other primary reasons may have had SARS-CoV-2 infection as a contributing, exacerbating factor. Additionally, this may not be discernable at the time of admissions, without further clinical workup and record review.

**eTable 4: Time since vaccination analysis: Rates per 100,000, Incidence Rate Ratio, and Vaccine Effectiveness**

| 5-11 | Jan 3 – 9 | Jan 10-16 | Jan 17-23 | Jan 24-30 |
|---|---|---|---|---|
| **Rates per 100,000** | | | | |
| Dec 13-19 | 379.6 | 259.1 | 164.0 | 99.1 |
| Dec 20-26 | 239.2 | 164.2 | 98.9 | 62.8 |
| Dec 27-Jan 2 | 168.9 | 123.8 | 75.9 | 48.4 |
| Unvaccinated | 484.0 | 256.3 | 130.7 | 70.0 |
| Dec 13-19 | 284.4 | 176.7 | 90.5 | 47.4 |
| Dec 20-26 | 233.1 | 120.3 | 61.1 | 42.6 |
| Dec 27-Jan 2 | 166.2 | 108.8 | 81.1 | 29.7 |
| Unvaccinated | 706.5 | 344.8 | 169.1 | 88.4 |
| **Incidence Rate Ratio** | | | | |
| **5-11** | | | | |
| Dec 13-19 | 1.27 | 0.99 | 0.80 | 0.71 |
| Dec 20-26 | 2.02 | 1.56 | 1.32 | 1.12 |
| Dec 27-Jan 2 | 2.86 | 2.07 | 1.72 | 1.45 |
| **12-17** | | | | |
| Dec 13-19 | 2.48 | 1.95 | 1.87 | 1.86 |
| Dec 20-26 | 3.03 | 2.87 | 2.77 | 2.08 |
| Dec 27-Jan 2 | 4.25 | 3.17 | 2.08 | 2.98 |
| **Vaccine Effectiveness** | | | | |
| **5-11** | | | | |
| Dec 13-19 | 21.6% | -1.1% | -25.5% | -41.5% |
| Dec 20-26 | 50.6% | 35.9% | 24.3% | 10.3% |
| Dec 27-Jan 2 | 65.1% | 51.7% | 41.9% | 30.9% |
| **12-17** | | | | |
| Dec 13-19 | 59.7% | 48.8% | 46.5% | 46.4% |
| Dec 20-26 | 67.0% | 65.1% | 63.9% | 51.8% |
| Dec 27-Jan 2 | 76.5% | 68.4% | 52.0% | 66.4% |
